# Genetic maternal effects contributes to the risk of Tourette’s disorder

**DOI:** 10.1101/2020.11.30.20240598

**Authors:** Behrang Mahjani, Lambertus Klei, Christina M. Hultman, Henrik Larsson, Sven Sandin, Bernie Devlin, Joseph D. Buxbaum, Dorothy E. Grice

## Abstract

**Background:** Risk for Tourette’s and related tic disorders (CTD) derives from a combination of genetic and environmental factors. While multiple studies have demonstrated the importance of direct additive genetic variation for CTD, little is known about the role of cross-generational transmission of genetic risks, such as maternal effects. Here, we partition sources of variation on CTD risk into direct additive genetic effect and maternal effects.

**Methods:** The study population consists of 2,522,677 individuals from the Swedish Medical Birth Register, born in Sweden between January 1, 1982, to December 31, 1990, and followed for a diagnosis of CTD through December 31, 2013.

**Results:** We identified 6,227 (0.25%) individuals in the birth cohort diagnosed with CTD. Using generalized linear mixed models, we estimated 4.7% (95% CrI, 4.4%-4.8%) genetic maternal effects, 0.5% (95% CrI, 0.2%-7%) environmental maternal effects, and 61% (95% CrI, 59%-63%) direct additive genetic effects. Around 1% of genetic maternal effects were due to maternal effects from the individual with comorbid obsessive-compulsive disorder.

**Conclusions:** Our results demonstrate genetic maternal effects contributing to the risk of CTD in offspring and also highlight new sources of overlapping risk between CTD and obsessive-compulsive disorder.

## 1. Introduction

Tic disorders are neurodevelopmental disorders that are diagnosed when motor and/or phonic tics onset during childhood and persist over at least one year. Tics typically wax and wane but are fundamentally involuntary behaviors whose severity and frequency can be influenced by other factors. A diagnosis of Tourette’s disorder, perhaps the most well-known of the tic disorders, requires the presence of both motor tics and at least one phonic tic. If only motor (or phonic) tics are present, the diagnosis of chronic motor (or phonic) tic disorder is given. Research has shown that Tourette’s disorder, chronic motor tic disorder and chronic phonic tic disorder can be considered phenotypic expressions of the same core genetic and biological underpinnings (1–3) and herein they are referred to as chronic tic disorders (CTD). The prevalence of CTD is estimated to be around 0.3-1% (4–6) and about half of the children with CTD are thought to be undiagnosed (7). CTD cluster in families at higher rates than expected by chance (8), consistent with shared genetic architecture and shared risk factors. Indeed, multiple studies demonstrated that 25%-77% of the liability of CTD is due to direct additive genetics (DG) (8–10).

Studies have linked maternal factors before and during pregnancy, *e*.*g*., maternal alcohol and cannabis use, inadequate maternal weight gain (11), and maternal smoking (12) to the risk of CTD. These conditions could represent maternal effects maternal effects. Maternal effects are influences on the offspring phenotype that result from maternal genotypes (above and beyond the genomic transmitted risk) or phenotypes. Maternal effects can be partitioned into genetic maternal effects (GME) and environmental maternal effects (EME). GME is a form of cross-generational transmission of genetic risk, and it occurs when the offspring’s phenotype is influenced by the genotype of the mother, independent of the offspring’s maternally inherited alleles. An EME occurs when the environment impacting the phenotype of the mother (independent of her genotype) influences the phenotype of the offspring. For the child, both GME and EME act as environmental effects. These categories of risk can be modeled and estimated by contrasting CTD risk in families of different relatedness.

Here, we investigate and test for the existence of maternal effects in the risk of CTD. We use a large population-based, prospectively ascertained cohort of Swedish-born individuals, and the relevant family data, to examine the role of DG, GME, and EME on the liability of CTD. We explore several models to adjust for potentially confounding factors such as sex, maternal psychiatric history, and paternal psychiatric history.

## 2. Method

### 2.1 Study population

The study population consists of all live-born singleton children born in Sweden between January 1, 1973, and December 31, 2000, with known father and mother as defined by the national Medical Birth Register (MBR) (13). Prospective follow-up continued until December 2013, and emigrated individuals identified during the follow-up were excluded from the study. To define family relationships, we included information about all relatives of each child using the Swedish Multi-Generation Registry (MGR)(14).

Ethical approval and waiver of informed consent were obtained from the Regional Ethical Review Board in Stockholm.

### 2.2 Outcomes

Diagnostic information about CTD was obtained using the Swedish National Patient Register (NPR), which includes diagnoses from inpatient and outpatient specialists (15). Sweden has a publicly financed healthcare system, and all visits to a specialist clinician are recorded with a diagnosis code using the International Classification of Diseases (ICD). All psychiatric hospital admissions in Sweden are recorded in the NPR from 1973. We used a validated diagnostic strategy developed by Rück *et al*. (16) to identify CTD cases based on the earliest registered CTD diagnosis code in the NPR (ICD-8: 306,2, ICD-9: 307C, and ICD-10: F95).

### 2.3 Exposure Covariates

We evaluated the following covariates for their relationship with CTD: sex of the child, maternal and/or paternal psychiatric history at the birth of the 1st child (yes/no for each). Maternal/paternal psychiatric history was defined as at least one psychiatric diagnosis for the mother/father (using ICD 7, 8, 9, or 10) at any time before the firstborn child.

### Statistical analysis

We used a similar approach applied in our previous study on the role of maternal effects on the risk of obsessive-compulsive disorder (OCD)(17). We defined family relationships based on first, second, and third-degree relatives: full siblings (FS), paternal and maternal half-siblings (pHS, mHS), and three different cousin types depending on whether the two parents responsible for the cousin relationship are sisters (maternal parallel cousins, mPC), bothers (pPC: paternal parallel cousins), or brother-sister (CC: cross cousins). A comparison of maternal and paternal half-siblings and cousins (mPC versus pPC and CC) delivers the required information to model and estimate maternal effects.

We estimated relative recurrence risks (RRR) of CTD for different relationship types using Cox proportional hazards regression, with attained age as the primary time scale and adjusted for sex and maternal and paternal psychiatric history. We bootstrapped families 1,000 times to estimate RRR (18). In the Cox regression, each individual was followed from 1997 until death, emigration from Sweden, diagnosis with CTD, or end of follow-up on December 31, 2013. We used generalized mixed linear models (GLMM) to partition the liability of CTD into covariates, DG, GME, EME, and individual variation (R). GME is a maternal cross-generational transmission of genetic risk and contributes to the risk of CTD in relatives with a maternal linkage, *e*.*g*., FS, mHS, and mPC. EME captures the effect of a shared environment that the biological mother provides for the offspring. Therefore, EME contributes to the risk of CTD in relatives with a common mother, *e*.*g*., FS, and mHS, while it is assumed to be zero for pHS and cousins (19)(Table S2).

We used a binary threshold-linear mixed model in a Bayesian framework with a non-informative prior to estimate the proportions of phenotypic variance explained by direct additive genetics and maternal effects (20). We applied a Gibbs sampler implemented in thrgibbs1f90b (20) and generated a sample size of 150,000, with 50,000 burn-in, from the posterior distribution of the variance components. Then, we calculated the mean of the posterior as the estimate of the variance components. The residual variance was fixed during the calculation. We reported the results with 95% Credible Intervals (CrIs), using Bayesian highest posterior density interval, which is analogous to two-sided 95% confidence intervals (CIs) in frequentist statistics (21). For the covariates, we reported the mean and standard deviation of the posterior to calculate the confidence intervals.

In the supplementary material, we used Falconer’s liability threshold model (LTM) for sensitivity analysis and compared the results with the estimates from GLMM.

## 3. Results

According to the ICD instruction manual and Diagnostic and Statistical Manual of Mental Disorders guidance, a diagnosis of transient tic disorder should be updated to a chronic tic disorder diagnosis if symptoms last more than one year. However, we had a few individuals with a diagnosis of transient tic lasting for more than one year. We removed all the individuals (n=367) that had a diagnosis of transient tic disorder at the end of the follow-up. For 20 of these individuals, the time between the first and the last diagnosis in NPR was more than one year. After cleaning the data, the cohort contained 2,522,677 individuals, of which 6.227 (0.25%) were diagnosed with CTD (48% female, Table 1). 967 of the parents in the cohort had a diagnosis of CTD.

**Table 1.** Study cohort including individuals born between 1982 and 1990.

|  | Total | Siblings |  |  | Cousins |  |  |
| --- | --- | --- | --- | --- | --- | --- | --- |
|  |  | Full | mHS | pHS | mPC | pPC | CC |
| **Participants** | 2,522,677 | 1,822,046 | 304,241 | 307,597 | 942,594 | 921,702 | 922,483 |
| **Male, count (%)** | 1,304,898<br>(52%) | 943,141<br>(52%) | 156,264<br>(51%) | 158,176<br>(51%) | 487,815<br>(52%) | 476,928<br>(52%) | 476,980<br>(52%) |
| **Female, count (%)** | 1,127,779<br>(48%) | 878,905<br>(48%) | 147,977<br>(49%) | 149,421<br>(49%) | 454,779<br>(48%) | 444,774<br>(48%) | 445,503<br>(48%) |
| **Diagnosed with CTD** | 6,227 | 3,957 | 1,128 | 1,035 | 2,075 | 2,083 | 2,219 |
| **Male, count (%)** | 4,843<br>(77%) | 3,092<br>(78%) | 916<br>(81%) | 797<br>(77%) | 1,612<br>(78%) | 1,621<br>(78%) | 1,723<br>(78%) |
| **Female, count (%)** | 1,384<br>(23%) | 865<br>(22%) | 212<br>(19%) | 238<br>(23%) | 463<br>(22%) | 462<br>(22%) | 496<br>(22%) |
| **Population Frequency** | 0.0025 | 0.0022 | 0.0037 | 0.0034 | 0.0022 | 0.0023 | 0.0024 |
| **Male** | 0.0037 | 0.0017 | 0.0030 | 0.0026 | 0.0017 | 0.0018 | 0.0019 |
| **Female** | 0.0011 | 0.0005 | 0.0007 | 0.0008 | 0.0005 | 0.0005 | 0.0005 |
| **Incidence rate\*** | 0.911 | 0.813 | 1.382 | 1.253 | 0.801 | 0.825 | 0.836 |

\*Per 10,000 person-years. mHS: maternal half-siblings, pHS: paternal half-siblings, mPC: maternal parallel cousins, pPC: paternal parallel cousins, CC: cross-cousins

We used Cox regression to compare the risk among different relation types (Table 2). Relative recurrence risk (RRR) for the full siblings was 17.586 (95% CI, 13.966-20.436). Maternal half-siblings had higher RRR than paternal half-siblings. RRR for maternal cousins (maternal parallel cousins) was higher than other cousins (paternal parallel cousins and cross cousins). Analysis of familial risk (FR) revealed a similar pattern (Table S1).

**Table 2.** Relative Recurrence Risk (RRR) for different relation types.

| Family Types | RRR | 95% CI |
| --- | --- | --- |
| Full siblings | 17.587 | (13.966-20.436) |
| Maternal half-siblings | 7.139 | (4.832-9.512) |
| Paternal half-siblings | 3.523 | (1.758-5.124) |
| Maternal parallel cousins | 2.099 | (1.273-2.813) |
| Paternal parallel cousins | 2.098 | (1.209-2.779) |
| Cross cousins | 1.927 | (1.175-2.490) |

We adjusted RRR for sex, maternal psychiatric history, and paternal psychiatric history (Table 3). RRR adjusted for maternal psychiatric history was smaller than RRR adjusted for paternal psychiatric history (16.634 vs. 17.120; Table 3).

**Table 3.** CTD: Adjusted Relative Recurrence Risk (RRR) for full siblings.

| Model | RRR | 95% CI |
| --- | --- | --- |
| Not Adjusted | 17.587 | (13.966-20.436) |
| Adjusted for Sex | 18.028 | (14.325-21.083) |
| Adjusted for Maternal Psychiatric History | 16.634 | (13.119-19.404) |
| Adjusted for Paternal Psychiatric History | 17.120 | (13.598-19.954) |

We used GLMM to estimate the contribution of DG and maternal effect on risk for CTD. The best GLMM model that explained the data was DG + GME + EME, as determined by Bayes Factor analyses (Table 4, and Table S5). 61% (95% CrI, 59%-63%) of the liability for CTD was due to DG (narrow-sense heritability), 4.7% (95% CrI, 4.4%-4.8%) due to GME, and 0.5 % (95% CrI, 0.2%-7%) due to EME. We used LTM to evaluate the sensitivity of the results (supplement, S1). The best model was 49% DG + 6.4% GME, suggesting that EME was zero or very small (Table S3).

**Table 4.**
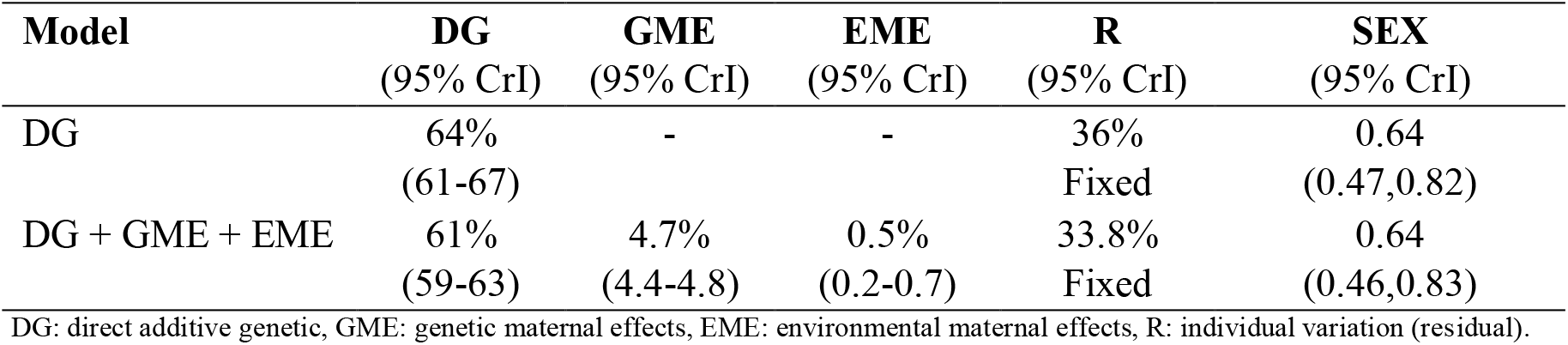
The proportion of phenotypic variance explained by different models.

| Model | DG<br>(95% CrI) | GME<br>(95% CrI) | EME<br>(95% CrI) | R<br>(95% CrI) | SEX<br>(95% CrI) |
| --- | --- | --- | --- | --- | --- |
| DG | 64%<br>(61-67) | - | - | 36%<br>Fixed | 0.64<br>(0.47,0.82) |
| DG + GME + EME | 61%<br>(59-63) | 4.7%<br>(4.4-4.8) | 0.5%<br>(0.2-0.7) | 33.8%<br>Fixed | 0.64<br>(0.46,0.83) |

DG: direct additive genetic, GME: genetic maternal effects, EME: environmental maternal effects, R: individual variation (residual).

Among the 6227 individuals with a diagnosis of CTD, 1012 were diagnosed with OCD. We removed the 1012 individuals with OCD from the cohort and estimated 47% DG and 5.1% GME using LTM (6.4% GME before removing OCD cases vs. 5.1% GME after removing OCD cases; Table S4). These results suggest that around 1.3% of the maternal effects for CTD was contributed by maternal effects for OCD.

## 4. Discussion

This is the first study that estimates genetic maternal effects as a cause of risk for CTD. Information for estimating maternal effects comes largely from the contrast of recurrence risk through maternal versus paternal lineages. The greatest information comes from maternal versus paternal half-sibling and cousin relationships, magnified when there is a multiplicity of affected siblings.

Using GLMM, and under the liability threshold assumption, we estimated 4.7% (95% CrI, 4.4%-4.8%) of the variance in risk is explained by GME, 0.5% (95% CrI, 0.2-0.7), and 61% (95% CrI, 59%-63%) by DG, while adjusting for the sex of the individual. Females were at 0.64 times higher risk, relative to males (95% CrI, 0.46-0.83).

The analysis of relative recurrence risk (RRR) of CTD was consistent with maternal effects in CTD risk architecture. RRR for maternal half-siblings was 7.139, while RRR for paternal half-siblings was 3.523; likewise, maternal parallel cousins versus other cousins carried a somewhat higher risk.

Previously we showed that genetic maternal effects accounted for 7.6% (95% CrI, 6.9%-8.3%) of the total variance in risk for OCD and direct additive genetics for 35% (95% CrI, 32.3%-36.9%). Since OCD and CTD share etiological overlap (8), we were interested in investigating if the observed maternal effects for CTD could be explained by maternal effects of the comorbid OCD cases. Using the weighted LTM model, we observed that around 1% of GME were due to maternal effects for the individual with OCD. This could be due to maternal pleiotropic effects or a shared genetic correlation. While the genetic correlation between CTD and OCD is reported to be around 0.41 (10), future studies are warranted to draw a more comprehensive picture of the genetic correlation, while accounting for shared maternal effects between these two disorders.

The present study had strengths and limitations. 1) In this study, we used data from the Swedish National Birth Register, which created a genetically homogeneous sample and would be expected to minimize the risk of confounding due to population stratification. 2) Our population study was based on clinical diagnoses of CTD, which would also be expected to reduce case misclassification and biases. 3) In theory, EME includes both shared and unshared environmental maternal effects (environmental maternal effects may be present for only some members of a sibship). However, the nature of the data and the statistical model used in this study captures only shared environmental effects. To estimate unshared environmental maternal effects, such as maternal illness during one pregnancy, individual information to that effect would need to be available. 4) The data are likely right-censored (some individuals may have received a diagnosis after study follow-up, and therefore these diagnoses would be missing from our data set), in particular for individuals born in the later years of the study. This would contribute to underdiagnoses of CTD and decreases the observed prevalence.

Risk for CTD derives from a complex combination of both nature (genome) and nurture (environment). Here, we demonstrated that maternal effects are directly causing risk and are not an effect of familial confounding factors. In addition, we showed that genetic maternal genetic (maternal genetic nurture (22)) plays a significant role in the risk of CTD.

## Data Availability

The data that support the findings of this study are available on request from the corresponding author, [DG]. The data are not publicly available.

## 5. Acknowledgments

This study was supported by a grant from the Mindich Child Health and Development Institute (DEG), the Friedman Brain Institute (DEG), and the Beatrice and Samuel A. Seaver Foundation (DEG, SS, JDB, BM), Icahn School of Medicine at Mount Sinai, New York, NY; the Mindworks Charitable Lead Trust (DEG); the Stanley Center for Psychiatric Research (DEG and JDB)

The sponsors of the study had no role in study design, data collection, data analysis, data interpretation, writing of the report, or in the decision to submit the paper for publication. The corresponding authors had full access to all data in the study and had final responsibility for the decision to submit for publication.

The computation was performed on resources provided by SNIC through Uppsala Multidisciplinary Center for Advanced Computational Science (UPPMAX) under Project SNIC 2016/1-359, SNIC 2017/7-113, and SNIC 2017/3-75.

## 6. Conflict of Interest

The authors report no biomedical financial interests or potential conflicts of interest.

## Notes

### Competing Interest Statement

The authors have declared no competing interest.

